# An online, open-access, cost comparison tool for presumptive tuberculosis testing using low complexity, automated, nucleic acid amplification tests (LCaNAAT) versus near point-of-care (NPOCs) molecular tests

**DOI:** 10.64898/2026.02.17.26346475

**Authors:** Pushpita Samina, Puneet Dewan, Chase Mertz, Mikashmi Kohli, Madhukar Pai

## Abstract

The world is far from achieving universal access to rapid tuberculosis (TB) diagnostics that align with the World Health Organization (WHO) TB diagnostic standard. Although WHO-recommended molecular rapid diagnostics (mWRDs), including low-complexity automated nucleic acid amplification tests (LCaNAAT), have significantly improved TB detection, scale-up has been constrained by high capital, maintenance, and consumable costs. Recently, lower-cost, near-point-of-care nucleic acid amplification tests (NPOCs) have emerged, with attractive global-access pricing, offering the potential to expand access to rapid TB diagnosis while reducing the financial burden. To support TB program staff better understand the budget impact of using newer NPOCs as the initial TB diagnostic compared with established mWRDs, we developed a flexible cost calculator. Originally built in Microsoft Excel for Mac (version 16.91, build 24111020), it organizes key parameters in a structured input sheet using standardized labels. Separate worksheets defined diagnostic scenarios and performed calculations, with results summarized in an overview covering various costs. After validation, the model was converted into R (version 2025.09.1 + 401). The app (app.R), developed with Shiny, is now available here for free global access. Using two hypothetical country case studies, we compared current LCaNAAT-based testing strategies with scenarios in which NPOCs were implemented as the initial diagnostic test. In hypothetical Country A, there were cost savings of 56.4% in Scenario 1 (based on current testing trend) and 60.5% in Scenario 2 (based on ambitious testing goals). In hypothetical Country B, the corresponding cost differences were 44.2% in Scenario 1 and 59% in Scenario 2. The analysis showed that countries can reach ambitious TB testing targets over 3 years using NPOCs at roughly the same total cost as implementing one year of current LCaNAAT testing. This highlights the potential of cheaper molecular diagnostics to significantly improve access to mWRDs. By reducing capital and maintenance costs, NPOC platforms allow TB programs to test and detect more cases well within existing budgets, speeding up progress toward universal rapid TB diagnosis.

## Introduction

The World Health Organization (WHO)-recommended molecular rapid diagnostic tests (mWRDs) - a broad class of WHO-endorsed nucleic acid amplification tests (NAATs) capable of simultaneous TB detection and drug susceptibility testing (DST) — and these were used as the initial test in fewer than half of newly diagnosed TB cases in 2024[1]. The most widely deployed platforms in this class are the Xpert MTB/RIF Ultra (Cepheid) and Truenat (Molbio), both low-complexity automated NAATs (LCaNAATs). To date, high maintenance costs, unsustainable donor-reliant funding models, and limited diagnostic access at the community level have obstructed the scale-up of mWRDs. While patients and providers consistently value fast, accurate, and affordable testing, TB programs often fail to meet these expectations [2]. Moreover, the sharp reduction in funding by the United States and other Group of Seven (G7) nations has introduced new uncertainty, threatening the End TB Strategy and the Global Plan 2023–2030 [3]. Even modest donor cutbacks could result in millions of additional TB cases and hundreds of thousands of preventable deaths by 2035 [4]. In this uncertain context, high burden countries will have to do more with less funding, invest more domestic resources, and be more self-reliant [5].

Thankfully, a new emerging class of simplified, portable molecular tests — near-point-of-care NAATs (NPOC-NAATs), hereafter referred to as NPOC platforms, (e.g., MiniDock MTB Test (MiniDock MTB))-have been developed for decentralized deployment at peripheral health facilities and community settings. Unlike LCaNAATs, NPOC platforms require minimal infrastructure, can accommodate non-sputum specimens such as tongue swabs, and are substantially lower in cost. Multicounty evaluations have shown that the NPOC achieves a sensitivity of 91.1% (95% CI: 82.1–95.9) and specificity exceeding 98% on sputum swabs [6], with WHO-pooled data across studies reporting a summary sensitivity of 85.4% (95% CI: 82.2–88.1) and specificity of 97.6% (95% CI: 96.9–98.1) on sputum. These estimates meet or approach the WHO Target Product Profile (TPP) thresholds of ≥90% sensitivity and ≥98% specificity for near-POC TB tests on sputum, and substantially outperform smear microscopy.

Importantly, with sputum swab-based testing, sensitivity for NPOC differed by only −3.0% (95% CI: −8.6, 2.6) compared to sputum Xpert Ultra, with specificity exceeding 98% [6] — a margin that is not statistically significant and supports the clinical equivalence of the two platforms in the populations studied. Based on the evidence, the WHO has recommended that, in adults and adolescents with signs and symptoms of pulmonary TB or who screen positive for pulmonary TB, NPOC on sputum should be used as the initial diagnostic test for TB rather than smear microscopy [7]. The product that met the performance criteria to establish the class of NPOC was the MTBC Nucleic Acid Test Card from Pluslife in Guangzhou, China. These test cards are designed to run on the Pluslife MiniDock amplification system instrument [7]. Further, the Global Drug Facility (GDF) has included the MiniDock technology at a price of $3.60 per test in its catalog [8]. Pluslife’s MTB Nucleic Acid Test Card is used on the Minidock Ultra device after sample pretreatment on the Thermolyse device; these devices are also available in the GDF Catalog for $155 and $180, respectively. These price points for tests and devices are substantially lower than the widely used mWRDs endorsed by WHO. However, the current version of the MiniDock test cannot provide results for rifampicin drug susceptibility. It is only designed for rapid TB detection.

To facilitate strategic adoption, estimate cost savings, and expand these new technologies, we created a free, simple cost-estimation tool. It initially launched as an Excel model and was later transformed into an interactive R web application. This tool assists national TB programs in assessing commodity costs and the budget implications of integrating NPOC platforms into current diagnostic networks. For example, if the National TB Control Program (NTP) of a high TB burden country were to switch from Xpert MTB/RIF Ultra to MiniDock as a first-line test for TB among people with presumed TB, what might the cost savings be for the country?

To address such pragmatic questions, our tool facilitates the estimation of the costs of testing presumptive TB cases using either NPOC devices (e.g., MiniDock) or low-complexity automated nucleic acid amplification tests (LCaNAATs), such as Xpert MTB/RIF Ultra. Designed to align with national budgeting and procurement cycles and the fiscal frameworks of global financing mechanisms such as the Global Fund, the tool serves as a simple, program-oriented budgeting resource. This paper describes the cost estimation tool, explains its underlying model, and provides guidance on its use. We have also developed two hypothetical country scenarios, Country A and Country B, modeled on publicly available data for Bangladesh and Indonesia, respectively, to elucidate the potential impact of NPOC use on diagnostic budgets. Its primary purpose is to help decision-makers understand the financial implications of expanding WRD access, optimizing resource use, and accelerating progress toward national TB control and case-finding targets. The purpose of our tool is not to do cost-effectiveness analyses on new TB tests.

## Materials and methodology

### Tool development

The initial version of the tool was created using Microsoft Excel for Mac (version 16.91, build 24111020). All relevant parameters were organized in an input sheet with consistent, clearly labeled variable names. Separate worksheets were used to define diagnostic scenarios and perform scenario-specific calculations, culminating in a consolidated output sheet that summarized results across multiple cost dimensions.

Once validated, the Excel-based tool’s underlying logic was translated into R (version 2025.09.1+401) to facilitate web deployment. The resulting application script (app.R) was developed using the Shiny package and deployed on a cloud-based platform (Fig 1) to provide interactive, free access to users worldwide (available here) (link: https://saminap.shinyapps.io/minidock/). The complete codebase is accessible here; link: https://zenodo.org/records/18653637.

**Fig 1:** Screenshot of the cloud-based cost estimating application.

### Scenarios modeled

To assess the financial implications of scaling up molecular TB testing, two programmatic scenarios were modeled to reflect varying levels of ambition and decentralization. The purpose was not to prescribe or recommend implementation strategies (since that will be a part of the upcoming WHO policy on NPOC tests), but rather to estimate how different scales of sample testing expansion could affect overall program costs. In each scenario, individuals with positive NPOC results are expected to have their specimens referred for reflex testing with an LCaNAAT for rifampicin resistance.

#### Scenario 1-current testing trend

This scenario represents the continuation of current diagnostic coverage and testing volumes, assuming the replacement of existing LCaNAAT testing with NPOC as the initial test for persons with presumptive TB, within existing operational capacity.

#### Scenario 2-ambitious testing goals

This scenario models an expanded, decentralized diagnostic network consistent with program-defined targets for scaling up WHO-recommended testing to all individuals with presumptive TB. It assumes increased test volumes and broader access through primary health care and community-based settings, which NPOC are capable of, but LCaNAAT are not. In this paper, for scenario 2, we assumed that all microscopy sites will be replaced by LCaNAAT and NPOC. For example, in Country A, in addition to existing sites, all microscopy sites (1,027) will be replaced with a new four-module LCaNAAT comparable to sites replaced by NPOCs.

While we selected these two for our description, within the tool, each scenario is fully customizable, allowing users to construct their own scenarios, and adjust structure and assumptions regarding testing targets, coverage, infrastructure, and cost components to fit specific country contexts.

### Time horizon

A three-year time horizon was applied to capture both one-time and recurrent costs. This approach allows inclusion of initial device purchase and installation costs, annual running and maintenance expenses, and potential reductions in pricing of emerging diagnostic products over time. By incorporating these dynamics, the model provides a more realistic estimate of the total programmatic cost of scaling up molecular testing under different implementation strategies.

### Tool structure

The application is organized into five functional modules:

1. Input module: Captures user-defined parameters, including health system capacity, TB epidemiology, diagnostic coverage targets, and cost data for devices and consumables. Default values are based on *ex-works* prices listed the Global Drug Facility (GDF) catalog and manufacturer price lists but can be edited to reflect country-specific procurement arrangements.
2. Intermediate outputs: Provide a disaggregated breakdown of cost categories, including equipment, consumables, maintenance, and sample transportation. For NPOC, additional costs related to reflex drug-resistant TB (DR-TB) sample testing are also calculated.
3. Final outputs: Summarizes annual and cumulative costs over three years for both diagnostic modalities. Differences between scenarios are calculated to enable direct comparison.
4. Visualizations: Includes automatically generated graphs depicting trends and comparative costs across the selected timeframe.
5. Documentation: Contains embedded formulae, definitions, and data source references to enhance transparency and replicability.

All modules are fully interactive, and inputs can be adjusted in real time to reflect changes in assumptions or programmatic strategies.

### Key assumptions

Table 1 summarizes the key assumptions for the model. Assumptions were established to simplify the model and make it user-friendly, based on publicly available data, discussions with local experts, and knowledge users.

**Table 1:**
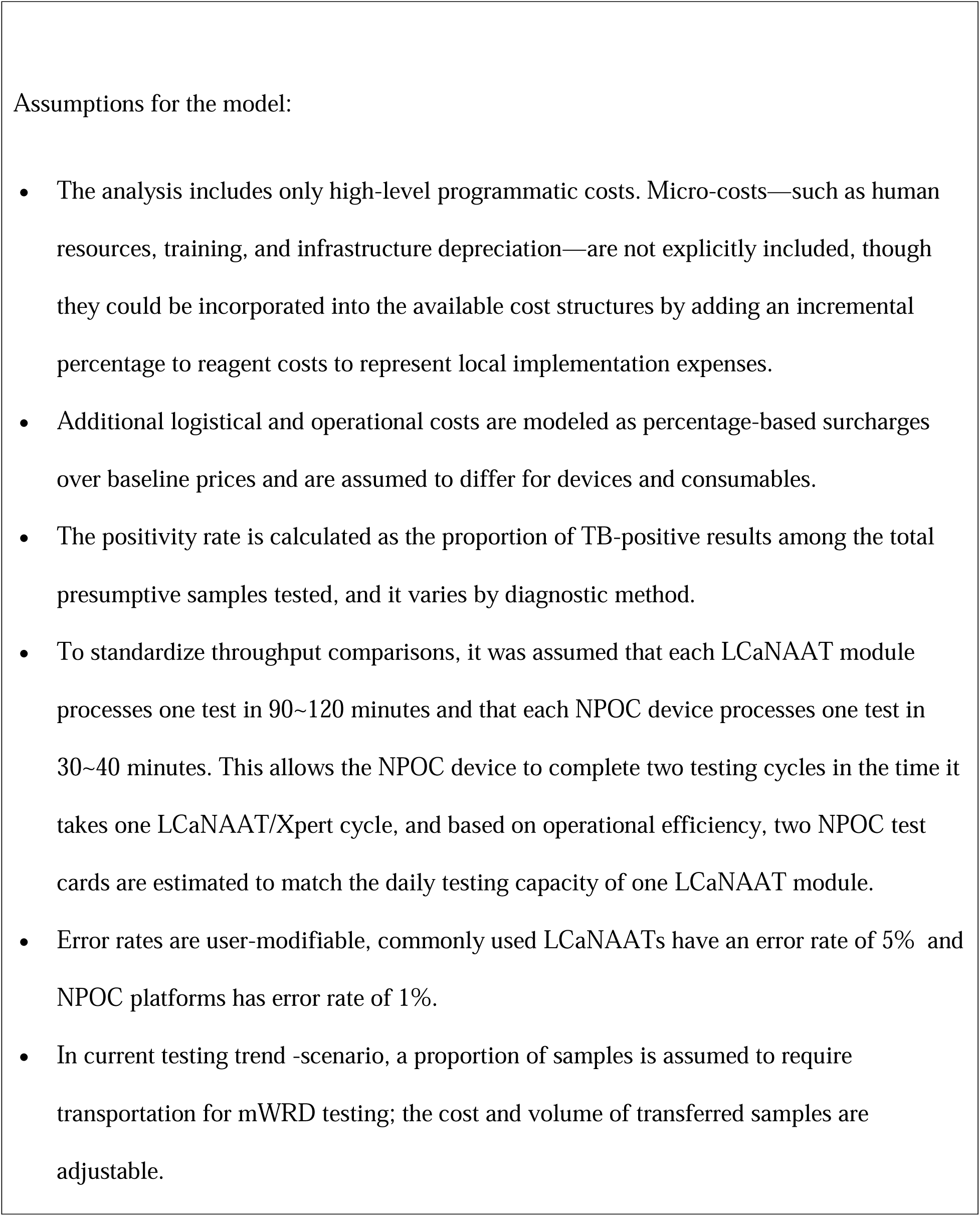
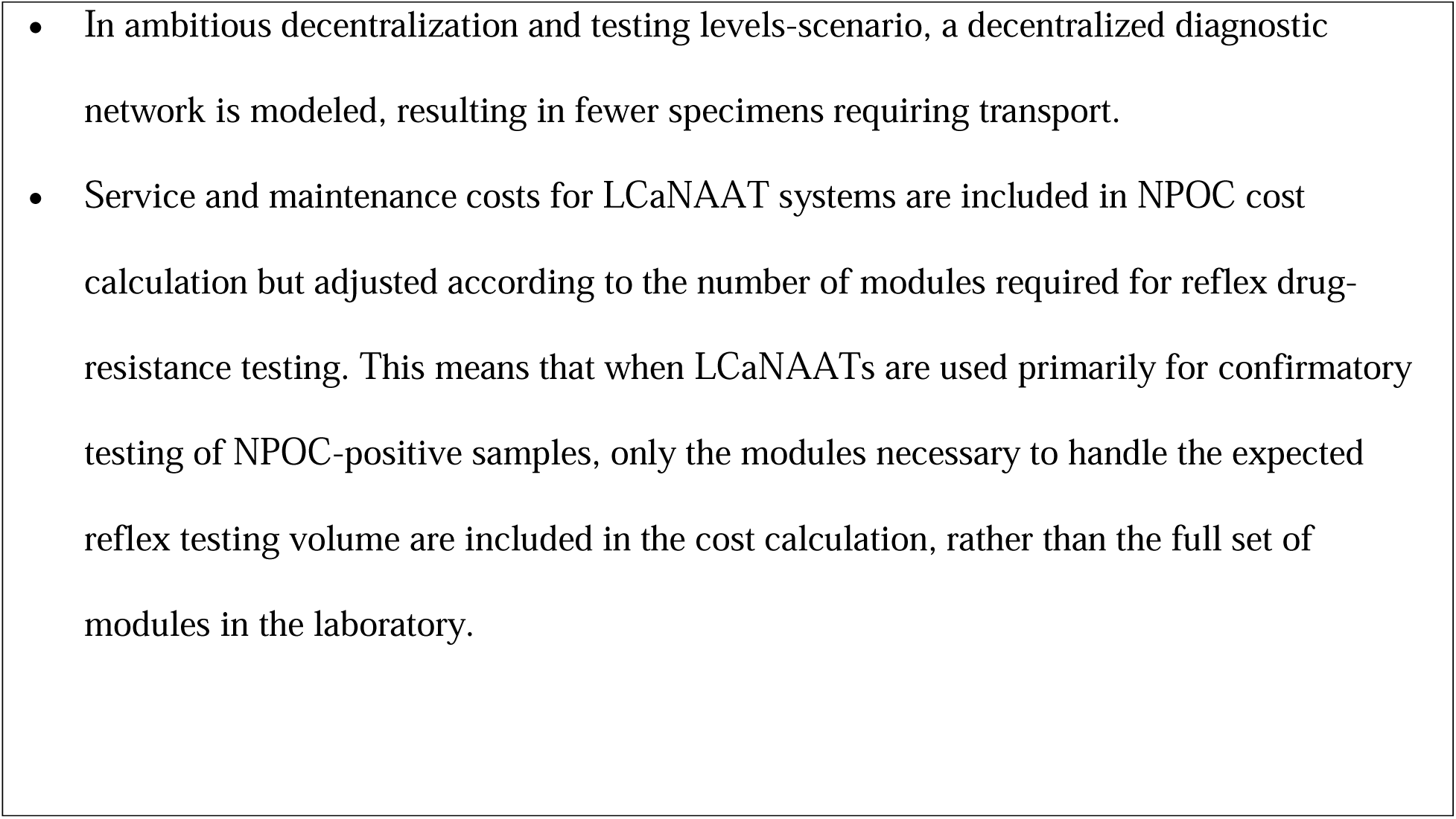
Key assumptions for cost comparison tool model.

### Input parameters

We utilized publicly available data and personal communications to model two hypothetical scenarios, Country A and Country B, which were selected to represent contrasting logistical and economic archetypes. Both countries expressed a specific interest in participating in this study, providing a unique opportunity to validate the model using real-world data from diverse settings:

Country A was chosen to represent a compact, high-density geography where decentralization is logistically streamlined (e.g., Bangladesh). It also models a favorable fiscal environment where diagnostic tools are exempt from customs duties.

Country B was selected to represent a vast, geographically fragmented setting where decentralization faces significant logistical hurdles (e.g. Indonesia). This scenario also accounts for a high-cost import environment, allowing the tool to demonstrate how trade barriers impact diagnostic scaling.

By using these two distinct profiles, the analysis illustrates how the benefits of decentralization vary across different national landscapes without relying on a single, uniform scenario.

### The epidemiological data and targets

#### Country A

Country A has an extensive network of LCaNAAT sites, comprising 4,416 LCaNAAT modules and 1,027 microscopy centers (numbers obtained from personal communication). In 2024, approximately 2.8 million individuals with presumptive TB were tested using mWRDs and microscopes, yielding 196,183 bacteriologically confirmed cases[1].

For scenario 1, we used only the molecular test targets, and for scenario 2, we used the targets for all tests (including microscopy and molecular tests) from 2028 to 2030, as specified in Country A’s National Strategic Plan (NSP) for 2026-2030.

Furthermore, based on personal communication, it was assumed that only 10% of total samples for presumptive tests were referred from microscopy sites to LCaNAAT facility. Considering the portability of NPOC and a more decentralized scenario, it was hypothesized that the implementation of NPOC would result in a reduction of sample transfers to the LCaNAAT site for primary testing to approximately 5% of the current volume under Scenario 1. The model estimates the number of reflex tests needing transfer to LCaNAAT sites for drug susceptibility testing (DST) based on the country’s implied positivity rates. Notably, diagnostic commodities procured through the Global Fund are currently exempt from customs and import duties in Country A. Nevertheless, a 2% charge for shipping and handling was incorporated into this analysis to account for associated logistical costs. Furthermore, storage and distribution costs were conservatively estimated at 2% of commodity value, in the absence of empirical data, consistent with assumptions applied in similar health economic analyses.

**Table 2:** Parameters & assumptions for Country A.

| Parameters | Assumptions |
| --- | --- |
| Total LCaNAAT modules/channels available | 4,416 |
| Number of microscopy sites | 1,027 |
| Presumptive TB tests done in a year | 2,800,000 |
| Total diagnosed TB cases (bacteriologically confirmed) | 196,183 |
| Implied Positivity rate (%) | 7 |
| Transportation costs (\$) | 5 |
| Percent of samples referred to molecular testing site (%) | 10 |
| Percent of samples referred to molecular testing site after NPOC rollout (%) | 5 |
| Approximate % import charge over baseline cartridge cost | 2 |
| Approximate % storage/distribution over baseline cartridge cost | 2 |
| Approximate % import charge over baseline device cost | 2 |
| Approximate % storage/distribution over baseline device cost | 2 |

**Table 3:** Testing target Inputs for Country A.

| Country A | Current testing trend |  |  | Ambitious testing targets |  |  |
| --- | --- | --- | --- | --- | --- | --- |
|  | Year 1 | Year 2 | Year 3 | Year 1 | Year 2 | Year 3 |
| Targets | 3,105,374 | 3,415,911 | 3,757,502 | 3,587,106 | 3,742,419 | 3,904,457 |

#### Country B

Country B has a substantial baseline diagnostic infrastructure, comprising 7,776 LCaNAAT modules and 7,927 microscopy testing sites that are not yet integrated with mWRDs. In 2024, approximately 4.7 million individuals with presumptive TB were tested using mWRDs and microscopes, yielding 804,836 bacteriologically confirmed cases[1].

For scenario 1, we used data from the WHO Global TB Report 2025 and assumed a 10% annual increase in these figures. This resulted in an estimated 3,709,525 molecular tests in the first year, with a 10% increase in each subsequent year. For scenario 2, we assumed all tests start at 5,634,577 in year 1 and increase by 10% per year. Based on communication with local experts, we expect 40% of samples to be transferred to molecular sites, which will decrease to 20% after the NPOC rollout in scenario 1, as NPOC-NAAT testing is possible at the lowest level of healthcare. Country B imposes customs duties on diagnostic cartridges and devices, including shipping and handling, which add approximately 20% to baseline costs. Storage and distribution costs are also higher than in Bangladesh, approximately 20% higher than baseline cartridge and device expenses.

**Table 4:** Parameters & assumptions for Country B.

| Parameters | Assumptions |
| --- | --- |
| Total LCaNAAT modules/channels available | 7,776 |
| Number of microscopy sites | 7,927 |
| Presumptive TB tests done in a year | 4,700,000 |
| Total diagnosed TB cases (bacteriologically confirmed) | 804,836 |
| Implied Positivity rate (%) | 17 |
| Transport cost (\$) | 5 |
| Percent of samples referred to molecular testing site (%) | 40 |
| Percent of samples referred to molecular testing site after NPOC rollout (%) | 20 |
| Approximate % import charge over baseline cartridge cost | 20 |
| Approximate % storage/distribution over baseline cartridge cost | 20 |
| Approximate % import charge over baseline device cost | 20 |
| Approximate % storage/distribution over baseline device cost | 20 |

**Table 5:** Testing Target Inputs for Country B.

| Country B | Current testing trend |  |  | Ambitious testing goals |  |  |
| --- | --- | --- | --- | --- | --- | --- |
|  | Year 1 | Year 2 | Year 3 | Year 1 | Year 2 | Year 3 |
| Targets | 3,709,525 | 4,080,477 | 4,488,524 | 5,634,577 | 6,198,034 | 6,817,837 |

### Cost parameters

LCaNAAT cost parameters are pre-populated using the Global Drug Facility (GDF) pricing catalog but may be customized by users to reflect negotiated rates, national tender agreements, or manufacturer-specific terms. In this model, for the device’s price and the installation cost, we have used the price of a four-module GeneXpert. Service costs were estimated by dividing the total device warranty by the module count, using a conservative baseline of USD 725. The baseline of USD 725 per module represents the estimated cost of a standard extended warranty covering basic parts and module replacement, providing a conservative lower bound relative to premium GDF Service Level Agreements like Warranty.

NPOC pricing is derived from the Minidock Pluslife catalog, with ex-works consumable costs set at USD 3.60 per test in Year 1 and decreasing to USD 2.50 in subsequent years. Devices are covered by a two-year warranty, with the option to purchase a Year 3 warranty extension. These costs can be adjusted in the tool.

**Table 2:** Cost parameters of LCaNAAT & NPOC.

| Cost | LCaNAAT (4 module) | NPOC | Source |
| --- | --- | --- | --- |
| Cost per device | \$ 19,000 | \$ 155 | GDF [8] |
| Installation cost | \$ 2,000 | \$400 | Assumption, GDF [8] |
| Specimen preparation device | N/A | \$ 180 | GDF [8] |
| Cost per test | \$ 7.97 | Year 1: \$ 3.6<br>Year 2 &3: \$ 2.5<sup>a</sup> | GDF [8], Assumption |
| Annual Service Warranty | \$ 725 per module | \$ 60 per device | GDF [8] |
| Error Rate (%) | 5 | 1 | [9] |

<sup>a</sup> It was assumed that the test cost would decrease as test volume increased.

Additional costs—such as packaging, shipping, which would increase duties, storage, and distribution—are included as user-defined percentages of baseline equipment and consumable costs. These allow for contextual adaptation to local procurement environments. Annual service charges are also modifiable to accommodate varying contractual or warranty arrangements.

Installation costs include laboratory setup, air conditioning, IT infrastructure, and related expenses—especially relevant in Scenario 2, where LCaNAAT platforms were introduced at a level equal to microscopy services.

#### Use case and adaptability

The tool is intended to inform national diagnostic strategies, particularly in low- and middle-income countries undergoing diagnostic scale-up or technology transition. By allowing users to adjust all relevant variables, the tool can be adapted to diverse settings with varying infrastructure, cost structures, and diagnostic priorities.

## Results

### Country A

#### Scenario 1: Current testing trend

Table 3 summarizes the results under both scenarios in Country A. Under Scenario 1, the total estimated cost of implementing LCaNAAT increased from USD 31.8 million in Year 1 to USD 37.8 million in Year 3. In contrast, NPOC implementation was substantially less expensive, ranging from USD 17.4 million in Year 1 to USD 14.6 million in Year 3.

The resulting annual cost differences between the two strategies grew over time, from USD 14.4 million in Year 1 to USD 23.2 million in Year 3. Across the 3 years, the cumulative savings from NPOC implementation instead of LCaNAAT was USD 58.8 million.

#### Scenario 2: More ambitious testing goals

In the scale-up scenario, we assumed that LCaNAAT modules or Nould will replace all microscopy sites,that and testing increased to meet the goals of the national strategic plan. With expanded testing, the cost gap between the two strategies widened further. The total annual cost of the LCaNAAT approach ranged from USD 42.6 million in Year 1 to USD 39.9 million in Year 3, while the corresponding NPOC implementation costs ranged from USD 19.5 million in Year 1 to USD 14.4 million in Year 3.

The annual cost difference increased from USD 23.1 million in Year 1 to USD 25.5 million in Year 3, demonstrating that cost savings associated with NPOC became more pronounced as testing volumes increased. Cumulatively over the 3 years, the NPOC implementation would save Country A USD 73.2 million. Table 3 (annex Tables 9 and 10) details the year-wise component costs.

**Table 3:** 3-Year Cost differences between LCaNAAT and NPOC in Country A (three-year cumulative cost in USD 2025).

| Costs | Scenario 1- Current testing trend |  | Scenario 2- Ambitious testing goals |  |
| --- | --- | --- | --- | --- |
|  | LCaNAAT only (USD) | NPOC+LCaNAAT (USD) | LCaNAAT only (USD) | NPOC+LCaNAAT (USD) |
| Total cost LCaNAAT test | 89,458,750 | 5,920,709 | 97,772,042 | 6,470,913 |
| Total cost NPOC test |  | 30,580,168 |  | 33,645,122 |
| Cost for additional devices |  | 1,687,795 | 5,592,320 | 2,080,697 |
| Service and maintenance | 9,604,800 | 1,064,905 | 11,840,700 | 1,568,847 |
| Sample transport | 5,139,394 | 6,141,208 | 5,616,991 | 3,903,408 |
| Total cost | 104,202,944 | 45,394,786 | 120,822,053 | 47,668,987 |
| Cost difference | 58,808,158 (56.4%) |  | 73,153,065 (60.5%) |  |

### Country B

#### Scenario 1: Current testing trend

Table 8 summarizes the Results of this scenario. Implementing only LCaNAAT results in testing costs that are consistently higher each year than the costs of implementing NPOC in Years 1-3. The analysis thus showed that there were significant savings from introducing NPOC, with savings increasing from $18.4. million in Year 1 to $33.1 million in Year 3. Overall, introducing NPOC in a scenario with a modest 10% annual increase in testing would cumulatively save Country B $82 million over the three years.

#### Scenario 2: More ambitious testing goals

Similar to Country A, in this larger scale-up of testing across Country B, the cost savings from implementing NPOC compared to using only LCaNAAT to replace all smear microscopy were even greater than the savings observed in the smaller scale-up of testing in Scenario 1. In Year 1, replacing all smear microscopy with NPOC and combining NPOC with LCaNAAT, rather than using LCaNAAT alone at the same scale-up, would save Country B over $90.4 million. In Year 3, NPOC implementation would save Country B more than $59.3 million. Overall, adopting NPOC and transitioning from LCaNAAT would cumulatively save Country B about $204.5 million over the 3 years.

**Table 4:** 3-Year Cost differences between LCaNAAT and NPOC in Country B (three-year cumulative cost in USD 2025).

| Costs | Scenario 1- Current testing trend |  | Scenario 2- Ambitious testing goals |  |
| --- | --- | --- | --- | --- |
|  | LCaNAAT only (USD) | NPOC+LCaNAAT (USD) | LCaNAAT only (USD) | NPOC+LCaNAAT (USD) |
| Total cost for test with LCaNAAT | 143,853,983 | 23,679,033 | 218,506,784 | 35,967,230 |
| Total cost for test with NPOC |  | 49,174,385 |  | 74,693,355 |
| Cost for additional NPOC devices |  | 4,000,752 | 56,685,200 | 8,079,708 |
| Service and maintenance | 16,912,800 | 3,622,962 | 34,156,200 | 7,288,173 |
| Sample transport | 24,557,052 | 22,889,313 | 37,300,896 | 16,117,239 |
| Total cost | 185,323,835 | 103,366,445 | 346,649,080 | 142,145,705 |
| Cost difference | 81,957,390 (44.2%) |  | 204,503,375 (59%) |  |

## Discussion

This analysis demonstrates that a simple, adaptable calculator can estimate three-year programmatic costs associated with NPOC uptake under both current and future testing goals. The tool is freely available as an open-access resource and can be customized to reflect any country’s diagnostic infrastructure, cost parameters, and strategic priorities. By allowing national TB programs to model different implementation scenarios, it provides a practical decision-support platform to inform planning and budgeting for molecular testing scale-up.

In our country examples, in Country A, the introduction of NPOC relative to continued exclusive use of LCaNAAT will result in an estimated cost reduction of approximately USD 58.8 million under current testing trends and USD 73.2 million under ambitious testing goals.

Similarly, in Country B (as an example), projected savings were approximately USD 82 million under current testing trends and USD 204.5 million under ambitious expansion. These results indicate that cost savings increase with testing scale, as the marginal cost of testing decreases once devices are in place and recurrent costs are limited mainly to consumables.

Notably, the analysis revealed that countries could implement ambitious national testing targets over 3 years using NPOCs at roughly the same total cost as maintaining one year of current LCaNAAT-based testing levels. This finding underscores the transformative potential of lower-cost molecular diagnostics to expand access to WHO-recommended rapid diagnostics (mWRDs). By reducing capital and maintenance costs, NPOCs can enable programs to test—and consequently detect—more presumptive TB cases within existing or only modestly increased budgets.

Beyond the direct cost savings, NPOCs offer operational advantages that strengthen their appeal for decentralized diagnostic networks. Their portability, low power requirements, and minimal installation needs make them suitable for peripheral or hard-to-reach health facilities, where LCaNAAT systems are often impractical. Decentralizing molecular testing to these levels could reduce specimen transport costs and diagnostic delays, improving patient access and accelerating case detection. Moreover, emerging evidence suggests that some NPOC platforms can process non-sputum samples such as saliva, further simplifying sample collection and enabling task-shifting to lower levels of the health system or private sector. Integrating NPOCs into community-based and private diagnostic networks could thus support a more equitable, people-centered approach to TB care.

A key additional consideration is that this analysis is a simplified, high-level costing tool and does not capture spatial or facility-level optimization of diagnostic networks. In practice, diagnostic network optimization analyses can provide important complementary insights by identifying optimal device placement, estimating realistic utilization rates, and determining required capacity across health system tiers. Such approaches could further refine investment planning and inform more detailed budget impact analyses by linking cost estimates to service delivery efficiency and geographic access.

However, NPOCs are not intended to fully replace LCaNAAT systems. For the time being, LCaNAAT platforms remain critical for drug-susceptibility testing (DST) and second-line diagnostic workflows. The complementary use of NPOC for initial screening and LCaNAAT for reflex DST testing offers a rational, tiered diagnostic approach that optimizes resource use while maintaining diagnostic accuracy and capacity for complex testing needs.

While these findings provide encouraging evidence for the economic potential of NPOCs, several caveats should be acknowledged. The cost estimates are based on modeled assumptions and represent a simplified view of program expenditures. The analysis focuses on major cost categories—equipment, consumables, service contracts, and sample transport—while excluding indirect and micro-costs such as training, human resources, supervision, infrastructure depreciation, and waste management. These elements may differ significantly across settings and could influence total implementation costs. Additionally, pricing assumptions reflect ex-works values and may not account for variations in import duties, supply chain inefficiencies, or local procurement practices.

Therefore, while the tool provides a useful high-level planning aid, it does not replace detailed cost-effectiveness or budget impact analyses. Future research should assess the real-world economic and clinical impacts of NPOC implementation, including patient-level outcomes such as time to diagnosis, treatment initiation, and overall TB detection rates. Field-based evaluations of operational feasibility, maintenance requirements, and user acceptability will also be critical for informing large-scale adoption.

Overall, this analysis highlights the promise of NPOC diagnostics as a cost-saving and scalable solution for expanding molecular testing coverage. It needs to be coupled with strategic investments in logistics and workforce capacity. NPOCs could play a pivotal role in helping countries meet national TB control targets and move closer to the WHO End TB Strategy goals.

## Data Availability

All codes are publicly available

https://zenodo.org/records/18653637

## Acknowledgement

We extend our sincere gratitude to Joshua Herbeck of the Gates Foundation, and Wayne Van Gemert of the Stop TB Partnership, as well as the National TB Control Program of Bangladesh and Indonesia, for their invaluable support and feedback during the development of this tool.

## Supporting information

S1 Fig. Fig 1 Screenshot of the cloud-based cost estimating application

S2 Table. Table 9 Scenario 1 cost components of LCaNAAT & NPOC in Country A

S3 Table. Table 10 Scenario 2 cost components of LCaNAAT & NPOC in Country A

S4 Table. Table 11 Scenario 1 cost components of LCaNAAT & NPOC in Country B

S5 Table. Table 12 Scenario 2 cost components of LCaNAAT & NPOC in Country B

